# A Novel Swarm Intelligence-Driven Feature Selection for Interpretable Machine Learning in GBM Overall Survival Analysis

**DOI:** 10.1101/2025.04.16.25325927

**Authors:** Abdulkerim Duman, Xianfang Sun, James R. Powell, Emiliano Spezi

## Abstract

**Purpose:** In this study, we develop and validate an interpretable machine learning (ML) model that integrates a hybrid Swarm Intelligence (SI)–based feature selection method with Magnetic Resonance Imaging (MRI)-derived radiomic features (RFs) to estimate overall survival (OS) in Glioblastoma Multiforme (GBM) patients. This study seeks to enhance the generalizability of the developed prognostic model and its potential for clinical integration by emphasizing feature reproducibility and leveraging multi-institutional retrospective datasets.

**Methods:** A cohort of 276 GBM patients with open-access pre-treatment MRI data (including T1, T1ce, T2, and FLAIR sequences) was used to perform comprehensive radiomic analysis. The extraction protocol yielded 1980 RFs per patient, extracted from three tumor regions (enhancing tumor: ET, tumor core: TC, and whole tumor: WT). The prognostic framework was built step-by-step, starting with a model of up to 10 RFs and then improving prediction by adding a single clinical feature (Age). In the training (discovery) dataset, we employed five-fold cross-validation combined with bootstrapping to ensure robust methodological validation. Model evaluation covered the C-index with 95% confidence intervals (CI) and survival stratification using Kaplan–Meier curves and the log-rank test to separate patients into low- and high-risk groups for OS.

**Results:** The final survival model integrates patient age and ten independent RFs; the model itself was optimized using features derived from three tumor contours and two MRI sequences (T1, FLAIR). The model’s performance in the holdout test dataset was evaluated by a concordance index (C-index) of 0.71 (95% CI: 0.61–0.79), exhibiting statistically significant risk stratification (p = 2 × 10⁻⁴). Upon external validation, the model achieved a C-index of 0.64, maintaining statistical significance (p = 1 × 10⁻²). The research combined the regularized Cox regression (Cox-LASSO), a traditional ML model, with a new SI-based LASSO-PSO method, yielding significant stratification. To our knowledge, the present study offers the first documented use of an interpretable ML model with an SI-based approach (LASSO-PSO) for successful risk stratification based on OS.

**Conclusion:** This study provides the development and validation of a clinical–radiomic model capable of conducting time-to-event analysis in GBM patients. By leveraging multicenter retrospective datasets, the model enables effective risk stratification based on OS. A key direction for future work involves exploring the combination of deep learning(DL)-based features and engineered features extracted via standardized convolutional filters, with the objective of improving OS prediction.

## 1. Introduction

Glioblastoma multiforme (GBM) is the most common primary brain tumor in adults, described by its rapid growth, aggressive nature, and significant infiltration of adjacent brain tissue [1], [2]. The World Health Organization (WHO) classifies GBM as a grade IV malignancy of the central nervous system. This classification represents the highest grade possible, highlighting the tumor’s extremely aggressive nature and the poor prognosis [3]. The aggressive invasion and rapid growth of GBM hinders successful surgical treatment, complicating surgical removal and undermining the effectiveness of conventional therapies [4]. GBM was predominantly diagnosed in individuals aged 75 to 84, revealing a significant gender difference of 5.6 cases per 100,000 among males compared to 3.5 in females [1].

GBM is characterized by intrinsic genetic complexity and pronounced heterogeneity at both intertumoral and intratumoral levels, which presents challenges in creating effective treatment options [4]. Biopsies are frequently used to diagnose GBM, which is important for effective treatment planning and reliable prognostic assessment [5]. Complementing conventional biopsies, non-invasive imaging modalities offer essential insights for the comprehensive characterization of tumors and their sub-regions [5]. In contrast to conventional imaging modalities such as computed tomography (CT) and positron emission tomography (PET), magnetic resonance imaging (MRI) provides better tissue contrast resolution and avoids exposing patients to ionizing radiation in healthcare [6].

As medical imaging analysis has progressed, radiomics has become a promising quantitative approach, allowing the extraction of computable features from MRI and CT scans [7], [8]. By providing quantitative insights into imaging patterns that are invisible to the human eye, radiomic analysis enhances the capacity for individualized treatment planning in GBM care, enabling more precise and effective therapeutic interventions.

Meneghetti et al. [9] adopted traditional approaches by utilizing 2 radiomic features (RFs), which yielded moderate predictive performance. Conversely, the guideline defined by van Timmeren et al. [10] declares that incorporating as many as 10 features optimizes the model’s effectiveness, offering a distinct perspective on methodological design.

Recent survival analyses in GBM integrate multiple clinical and RFs, although the inclusion of numerous features can limit reproducibility and interpretability [11], [12], [13]. In radiomics, swarm intelligence (SI) methods, most notably Genetic Algorithms (GA) and Particle Swarm Optimization (PSO), are increasingly employed for feature selection, presenting promising solutions for analyzing high-dimensional data [14], [15], [16], [17]. Hybrid methods that utilize SI algorithms might yield effective solutions for feature selection and predictive modeling in radiomics, demonstrating competitive or superior performance while maintaining interpretability in applications related to GBM.

This research proposes a new hybrid feature selection framework that merges elements from Meneghetti et al. [9] and Al-Tashi et al’s [15], utilizing an SI-supported LASSO to improve the accuracy and efficiency of feature selection in machine learning (ML) applications. To address reproducibility challenges in radiomics-based GBM survival analysis, this study utilizes widely used preprocessing and segmentation methods, validating its findings with open-access data from multiple institutions. This research is the first to use an SI-based feature selection with traditional ML algorithms for GBM time-to-event radiomic analysis, focusing on their effectiveness in stratifying patient risk.

## 2. Material and Methods

This study employed a radiomic model development and validation framework based on 276 GBM cases, integrating data from two sources: 236 cases from the multi-institutional BraTS 2020 Challenge dataset [18], [19], [20], and 40 cases from the single-institutional Río Hortega University Hospital Glioblastoma Dataset (RHUH-GBM) [21]. In compliance with the Response Assessment in Neuro-Oncology (RANO) criteria [22], both datasets comprised four standardized preoperative MRI modalities: T1-weighted (T1), T1-weighted contrast-enhanced (T1ce), T2-weighted (T2), and Fluid-attenuated inversion recovery (FLAIR). Moreover, they included information on overall survival (OS) and patient age. The study, as shown in Figure 1, applied a time-to-event analysis for OS, stratifying cases into high- and low-risk groups. The endpoint was measured from the date of pathological diagnosis to death (censored = 1) or final follow-up (censored = 0). As part of a rigorous study design, the BraTS 2020 dataset was divided into two subsets. The discovery cohort, representing 80% of the data (n = 188), was utilized for model development and internal validation processes (n = 48). Additionally, the RHUH-GBM dataset was employed as an external validation set to evaluate the generalizability of the model. Clinical parameters from the discovery cohort were combined with RFs extracted from three tumor regions: enhancing tumor (ET), tumor core (TC), and whole tumor (WT). Feature selection was conducted, followed by the development and optimization of risk stratification models, utilizing the discovery cohort for analysis. Model performance was evaluated using a two-step validation strategy: first, by testing on a hold-out dataset, and second, by applying the models to an external validation cohort to assess their generalizability.

**Figure 1.**
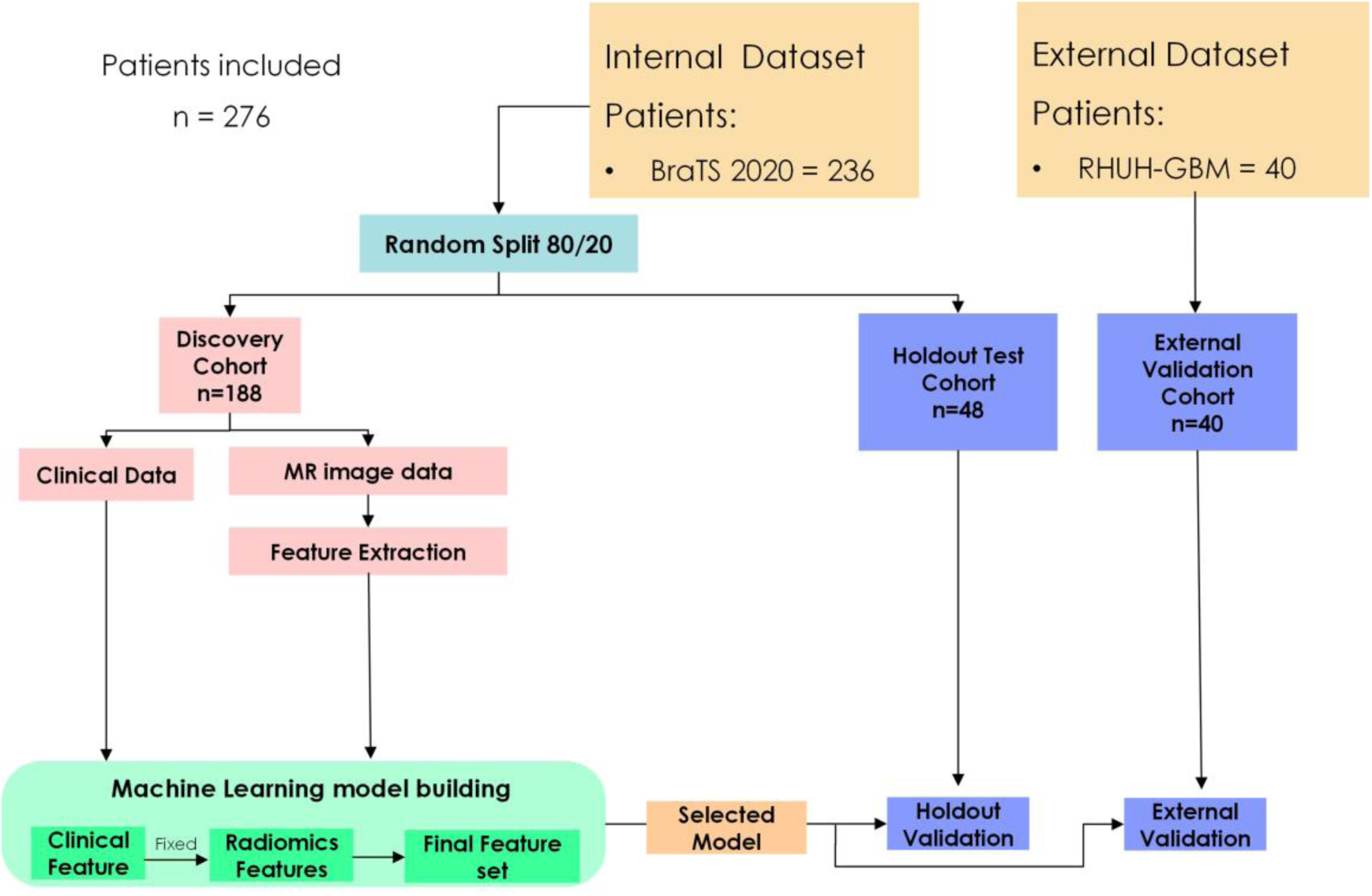
This study used an internal dataset, which consisted of a discovery cohort for selecting features and developing the model, along with a holdout test cohort for assessing performance on unseen data. Additionally, an external dataset was utilized to further validate the model’s generalizability.

Risk stratification performance was assessed using the concordance index (C-index). Kaplan-Meier plots and log-rank tests were utilized to evaluate differences in survival outcomes across risk groups.

The BraTS dataset incorporates MRI scans from 19 institutions, representing a broad spectrum of imaging protocols and scanner settings. Preprocessing included DICOM-to-NIfTI conversion, followed by N4 bias field correction as a preliminary step for registration [23]. The spatial alignment procedure involved a series of steps: first, aligning the T1, T2, and FLAIR sequences to the T1ce volume, and then registering them to the SRI24 atlas framework [24]. Consistent spatial sampling was achieved through isotropic voxel resampling (1 × 1 × 1 mm³). A skull-stripping method was carried out using a pre-trained DL model. Z-score normalization was applied to mitigate scanner- and patient-specific intensity variations. Using CaPTk [25], the preprocessing pipeline included brain tissue extraction, intensity standardization, and spatial normalization, resulting in MRI volumes standardized to 1 × 1 × 1 mm³ voxel resolution and 240 × 240 × 155 dimensions.

To ensure compatibility with the BraTS 2020 dataset, the RHUH-GBM dataset was processed using a consistent preprocessing pipeline [21]. This included: (1) DICOM-to-NIfTI conversion; (2) rigid registration of T1ce images to the SRI24 atlas, with the subsequent alignment of T1, T2, and FLAIR sequences to the transformed T1ce volume; (3) brain extraction performed on all registered sequences using a DL-based method; and (4) Z-score intensity normalization. Uniformity in voxel resolution (1 × 1 × 1 mm³) and matrix dimensions (240 × 240 × 155) was preserved across all scans.

In accordance with IBSI guidelines [26], image preprocessing was conducted to support reproducibility. A two-phase approach was employed for tumor segmentation in the BraTS 2020 and RHUH-GBM datasets: automatic segmentation using DL techniques, followed by confirmation from neuroradiologists to ensure compliance with clinical standards [18], [21]. For each patient, 1,980 RFs were extracted, derived from a combination of four MRI modalities and three tumor regions, with 165 features computed per region (4 × 3 × 165). The extraction process was conducted using the MATLAB-based version of the Spaarc Pipeline for Automated Analysis and Radiomics Computing (SPAARC, https://www.spaarc-radiomics.io/, accessed on 1 January 2025) [27], [28]. Tumor characteristics such as shape, texture, and intensity were quantified through radiomic feature extraction. To promote reproducibility and consistency across datasets, all features were derived using a three-dimensional (3D) methodology and standardized in compliance with IBSI guidelines. Details of the preprocessing parameters and extracted RFs are presented in Figure S-1.

To promote methodological rigor and reduce overfitting, model development followed a structured pipeline that employed three complementary feature selection approaches. The modeling framework (Figure 2), consisted of four stages: (i) feature preprocessing, (ii) feature selection, detailed in Figure S-3 (LASSO-RANK), Figures S-4 and Figure S-5 (LASSO-GA), and Figures S-6 and Figure S-7 (LASSO-PSO), (iii) hyperparameter tuning, and (iv) model training with internal validation on the discovery cohort. All steps were conducted using five-fold cross-validation.

i. To address feature scale variability, Z-score normalization was conducted on the training (discovery) dataset. The resulting parameters were then used to normalize the holdout test and external validation datasets.
ii. The feature selection methods cover three methods: one traditional approach and two novel variations that employ algorithms based on utilizing SI-based feature selection algorithms [29]. The established LASSO-RANK algorithm [30], outlined in Leger et al. [31], provided the fundamentals. Furthermore, this study proposed a novel two-phase hybrid feature selection method. The first step was a key modification on LASSO-RANK, which excluded only the frequency-based feature ranking step after generating a feature pool. This pool was subsequently utilized using SI algorithms, specifically GA [32] and PSO [33], to enhance the feature selection step. The proposed models are LASSO-GA and LASSO-PSO. The LASSO-RANK method, used as the baseline approach, employed a five-fold cross-validation strategy to identify a subset of up to nine RFs, ranging from 2-pair to 9-pair RFs. In the novel two-phase approach, either GA or PSO was used to refine the feature pool obtained from LASSO regression. In contrast to LASSO-RANK’s deterministic feature selection, GA and PSO apply stochastic (random), iterative search strategies to optimize feature subset selection. The hyperparameter configurations for these algorithms are provided in Figure S-2. LASSO was the estimator for algorithms, with a negative mean squared error metric for each SI-based feature selection step. Risk stratification was conducted utilizing two survival analysis models: regularized Cox regression (Cox-LASSO) [30] and Random Survival Forests (RSF) [34]. These frequently used models [35] have been tailored for survival analysis, optimizing to effectively handle censored time-to-event data.
iii. Model hyperparameters were tuned via bootstrap resampling on the discovery cohort to minimize overfitting and improve the model’s risk-stratification performance on unseen data.
iv. Following the guidelines of van Timmeren et al. [10], the total feature number, including patient age as a clinical variable, was limited to ten. LASSO-RANK identified 2 to 9 radiomic feature pairs per fold through five-fold cross-validation. The eight feature subsets, ranging from 2 to 9 features, were then ranked within the aggregated eight feature pools based on selection frequency. To assess risk stratification performance, model validation was performed using 200 bootstrap resampling iterations on the discovery cohort with the selected feature subsets. Additionally, LASSO-GA and LASSO-PSO also utilized the eight feature pools to select the final feature subsets, applying the same model validation strategy.

**Figure 2.**
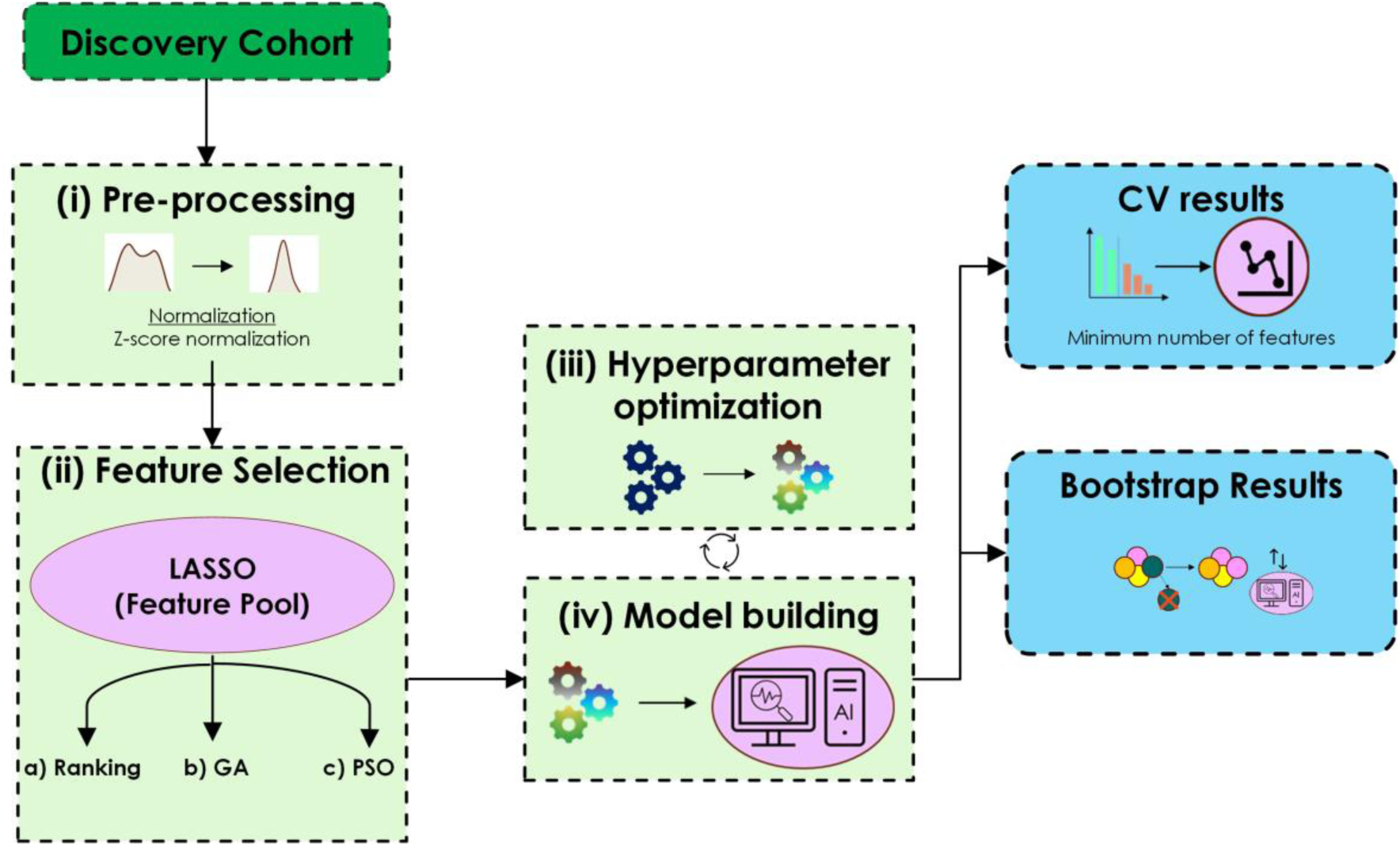
The radiomic workflow for Feature Selection methods and Hyper-parameter optimization.

The C-index was used as the primary performance metric to evaluate robustness and predictive accuracy, with model development and optimization conducted on the discovery cohort. Performance evaluation was carried out on two datasets: the holdout portion of BraTS 2020 and an external validation set (RHUH-GBM). By implementing a two-stage validation, the study aimed to assess both model performance and generalizability. Survival distributions were compared using the log-rank test in the discovery and both the holdout test set and the external validation cohort. To assess statistical differences in continuous variables, the Mann-Whitney U test was applied between the discovery cohort and both the holdout test set and the external validation cohort. The Kaplan–Meier curve was used to assess risk stratification, with patients divided into low- and high-risk groups according to the median risk score as the cut-off. To determine the statistical difference in survival outcomes between high- and low-risk groups, the log-rank test was employed. The prognostic models’ stratification performance was measured using the C-index, with 200 bootstrap iterations performed across the discovery, holdout test, and external validation cohorts to estimate the 95% Confidence Interval (CI)) [36]. All statistical and survival analyses were performed in Python v3.10, with p < 0.05 considered statistically significant. Image preprocessing and analysis steps are outlined in Figure 2. Permutation feature importance was used for feature relevance assessment via Scikit-learn v1.5.2. PSO and GA were implemented using ps-optimize v2.0.4 and sklearn-genetic v0.6.0, respectively.

## 3. Results

Table 1 summarizes the clinical characteristics of the discovery, holdout test, and external validation cohorts. The discovery cohort presented a median OS of 12.05 months, while the hold-out test cohort exhibited a median OS of 14.44 months. Statistical analysis indicated that the difference in median OS between these cohorts was not significant (p = 0.58). The external validation cohort had a median OS of 12.13 months, which was not significantly different from that of the discovery cohort (p = 0.59). Analysis indicated that the LASSO-RANK and LASSO-GA methods achieved peak performance with a six-pair feature pool. In comparison, LASSO-PSO benefited from larger feature sets, requiring a nine-pair pool to reach its optimal performance level. To address multicollinearity, an initial step was carried out before the feature selection process. To reduce feature redundancy, Spearman correlation coefficients were calculated across all RFs. Subsequently, features demonstrating high correlation (ρ > 0.95) were excluded, yielding a refined feature set of 767 RFs. The final feature pools were constructed by aggregating RFs selected across all five cross-validation folds, with repetitions excluded. This process yielded 16 RFs for both LASSO-RANK and LASSO-GA (each from a six-pair pool) and 18 RFs for LASSO-PSO (from a nine-pair pool). For the LASSO-RANK method, the final model utilized the six features demonstrating the highest selection frequency, as the best performance was previously achieved with a configuration of the six-pair feature pool.

**Table 1.** Characteristics of clinical variables for discovery and hold-out test and external validation datasets.

| Variable | Discovery Dataset<br>Median<br>(range) | Holdout Test Dataset<br>Median<br>(range) | Statistical Cohort Comparison | Discovery Dataset<br>Median<br>(range) | External Validation Dataset<br>Median<br>(range) | Statistical Cohort Comparison |
| --- | --- | --- | --- | --- | --- | --- |
| Age (Years) | 62.4 [19.0-86.7] | 60.6 [27.8-85.9] | U: 0.55,<br>p-value: 0.3 | 62.4 [19.0-86.7] | 64.0 [45.0-78.0] | U: 0.46,<br>p-value: 0.55 |
| OS (months) | 12.05 [0.17-52.03] | 14.44 [1.0-58.9] | U: 0.47,<br>p-value: 0.58 | 12.05 [0.17-52.03] | 12.13 [3.0-41.47] | U: 0.47,<br>p-value: 0.59 |

Internal validation guided the final selection of feature sets for LASSO-GA and LASSO-PSO, which included 2 and 10 RFs, respectively, as shown in Table 2. Cox-LASSO and RSF hyperparameters were tuned using 200 bootstrap iterations on the full discovery cohort, based on the selected RFs and patient age as the clinical feature. Final hyperparameter configurations were optimized for each model-feature selection method combining using k-fold cross-validation and bootstrap techniques; all hyperparameter configurations are presented in Supplementary Materials Table S-1. The development of prognostic models included the full discovery cohort, combining patient age info with the selected RFs to create an integrated clinical-radiomic signature. Through the application of LASSO-based feature selection techniques, a subset of robust RFs was identified across diverse MRI sequences, preserving performance with fewer features.

**Table 2.**
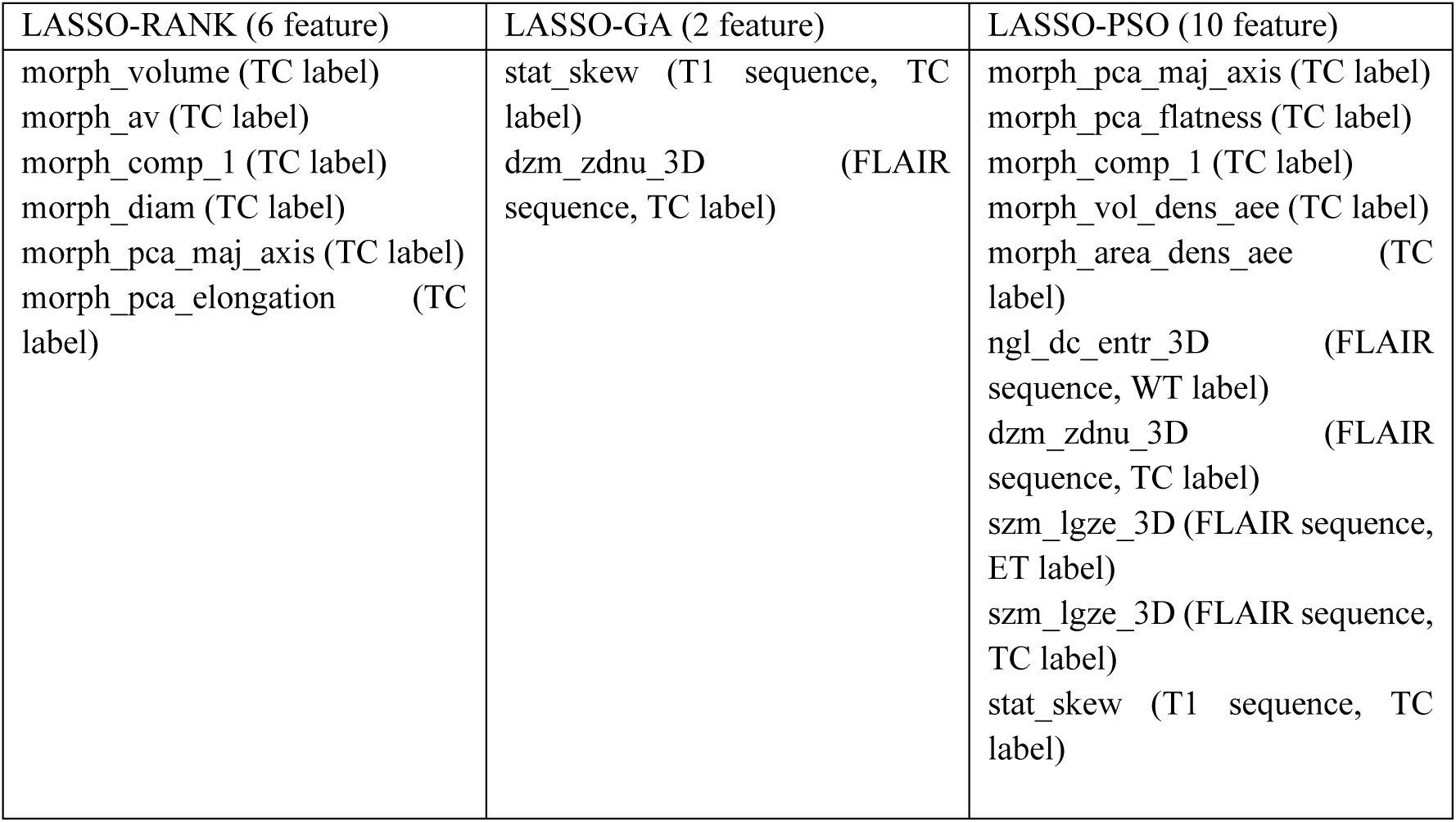
The selected RFs for each feature selection method, with their respective MRI sequences and Labels indicated in parentheses. Notably, Morphological features are associated with contour morphology (shape) rather than a specific MRI sequence. The MRI sequences used for radiomic feature extraction are specified in parentheses (FLAIR sequence, T1 sequence), along with the regions of interest (ROIs) utilized for feature extraction (e.g., ET label, TC label, WT label).

| LASSO-RANK (6 feature) | LASSO-GA (2 feature) | LASSO-PSO (10 feature) |
| --- | --- | --- |
| morph_volume (TC label)<br>morph_av (TC label)<br>morph_comp_1 (TC label)<br>morph_diam (TC label)<br>morph_pca_maj_axis (TC label)<br>morph_pca_elongation (TC label) | stat_skew (T1 sequence, TC label)<br>dzm_zdnu_3D (FLAIR sequence, TC label) | morph_pca_maj_axis (TC label)<br>morph_pca_flatness (TC label)<br>morph_comp_1 (TC label)<br>morph_vol_dens_aee (TC label)<br>morph_area_dens_aee (TC label)<br>ngl_dc_entr_3D (FLAIR sequence, WT label)<br>dzm_zdnu_3D (FLAIR sequence, TC label)<br>szm_lgze_3D (FLAIR sequence, ET label)<br>szm_lgze_3D (FLAIR sequence, TC label)<br>stat_skew (T1 sequence, TC label) |

Identified for its high interpretability [37], the Cox-LASSO model achieved the highest C-index of 0.64 on internal validation when using features selected by LASSO-PSO or LASSO-GA (Figure 3b). According to the out-of-bag (OOB) bootstrap evaluation on the discovery dataset (Figure 3c), the prognostic model achieved a C-index of 0.67 using LASSO-PSO selected features, outperforming the model with LASSO-GA (C-index = 0.65), representing greater robustness with LASSO-PSO selection. Internal validation of the LASSO-PSO RSF models showed consistent internal validation results, yielding a C-index of 0.64 (Figure 3b). Despite demonstrating strong OOB bootstrap performance with a C-index of 0.74, the best-performing RSF model using LASSO-PSO performed limited generalizability to unseen datasets (Supplementary Materials Table S-2).

**Figure 3.**
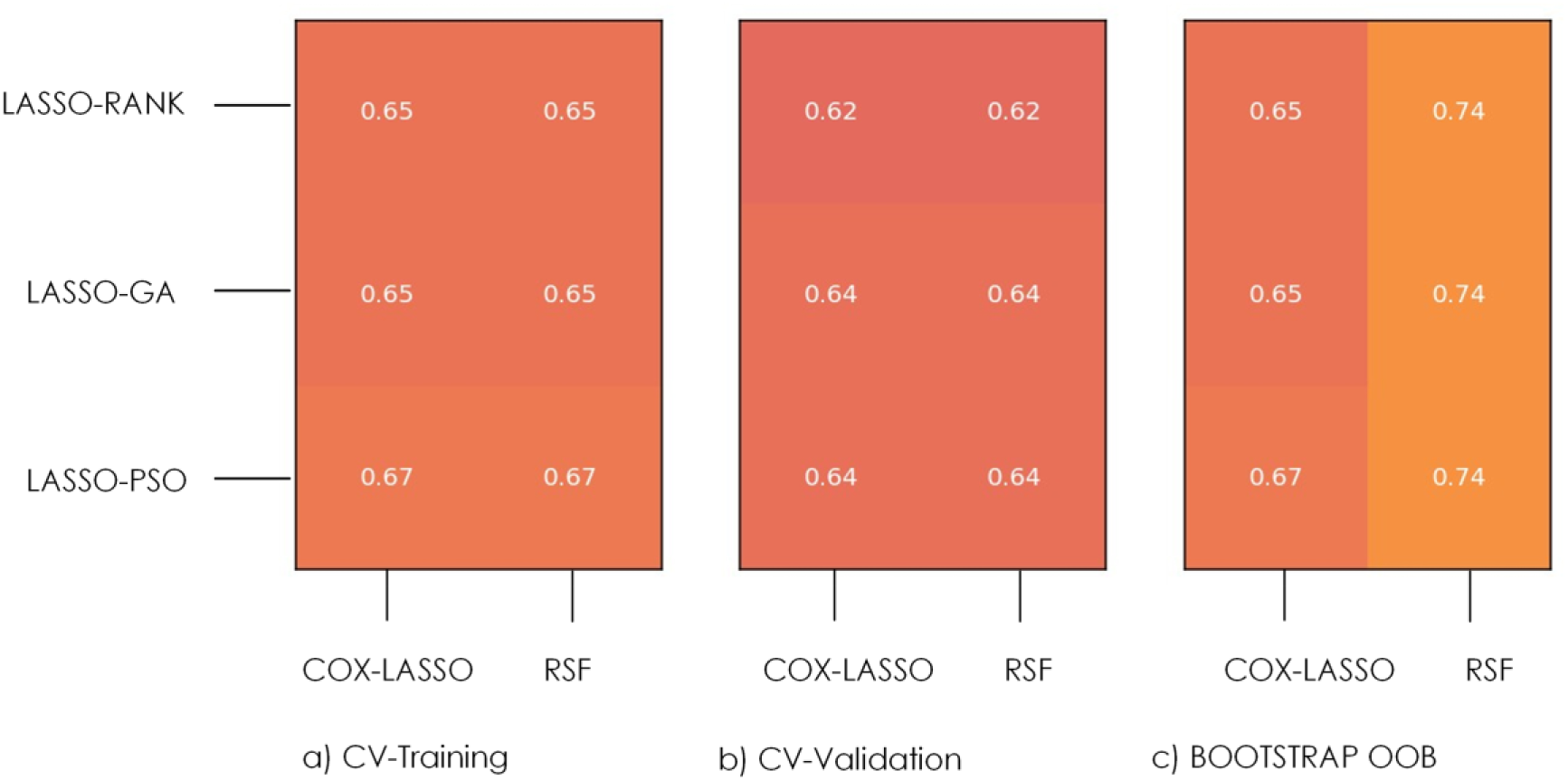
The C-index values for each model on each feature selection method and their corresponding ML algorithms were assessed for GBM time-to-event analysis. The results are presented as follows: (a) Cross-validation (CV) training performance, (b) Cross-validation (CV) validation performance, and (c) Bootstrap out-of-bag (OOB) evaluation.

The Cox-LASSO model was built using 10 RFs (Table 2) within the discovery cohort exhibited notable predictive accuracy, as reflected by a C-index of 0.64 with a 95% CI of 0.60–0.68. The texture feature Dzm_zdnu_3D, using TC label, had the highest hazard ratio (HR = 1.15) in the 10-feature radiomic model (Table 3), with a 95% CI of 0.87–1.76. This feature quantifies the distribution uniformity of zone frequencies through spatial distances. Low scores reflect zone homogeneity in the tumor; high scores reflect clustering and greater intertumoral heterogeneity. The selected feature set comprised an equal distribution between MRI sequence-dependent and shape-based features: 50% were morphological (shape-based, 5/10) features not tied to a specific MRI sequence, and the remaining 50% were extracted from FLAIR (4/10) and T1 (1/10) sequences. These RFs exhibited low inter-correlation (Spearman’s ρ < 0.3), suggesting independent contributions. On the hold-out test dataset, the radiomic model achieved a C-index of 0.61 (95% CI: 0.52–0.72), demonstrating moderate prognostic performance in Table 3. Morph_comp_1, a shape-based feature, emerged as the most notable predictor, exhibiting the highest hazard ratio (HR) of 2.81 (95% CI: 0.50–15.15), as documented in Table 3. By evaluating the geometrical conformity of the targeted ROI to a perfect spherical morphology, this feature serves as a metric for characterizing tumor morphological compactness.

**Table 3.** Univariate (Clinical Model) and Multivariate (Radiomic Model) Cox regression analysis for Discovery and Holdout test datasets.

|  |  | Discovery cohort | Holdout test cohort | Discovery cohort |  | Holdout test cohort |  |
| --- | --- | --- | --- | --- | --- | --- | --- |
| Model | Variable | HR [95% CI] | HR [95% CI] | p-Value | C-Index | p-Value | C-Index |
| Clinical Model | Age | 1.32 [1.16-1.52] | 1.78 [1.44-2.58] | $3 \times 10^{-2}$ | 0.59 [0.55-0.64] | $7 \times 10^{-5}$ | 0.71 [0.62-0.79] |
| Radiomic Model | morph_pca_maj_axis (TC label) | 1.08 [0.77-1.47] | 2.06 [0.85-8.72] | $4 \times 10^{-4}$ | 0.64 [0.60-0.68] | $2 \times 10^{-2}$ | 0.61 [0.52-0.72] |
|  | morph_pca_flatness (TC label) | 1.13 [0.81, 1.42] | 1.39 [0.62, 3.74] |  |  |  |  |
|  | morph_comp_1 (TC label) | 0.92 [0.57, 1.51] | 2.81 [0.50, 15.15] |  |  |  |  |
|  | morph_vol_dens_aee (TC label) | 0.89 [0.55, 1.45] | 0.41 [0.07, 2.13] |  |  |  |  |
|  | morph_area_dens_aee (TC label) | 1.12 [0.83, 1.70] | 2.38 [0.26, 12.77] |  |  |  |  |
|  | ngl_dc_entr_3D (FLAIR sequence, WT label) | 0.91 [0.76, 1.07] | 0.41 [0.15, 0.84] |  |  |  |  |
|  | dzm_zdnu_3D (FLAIR sequence, TC label) | 1.15 [0.87, 1.76] | 1.14 [0.56, 2.51] |  |  |  |  |
|  | szm_lgze_3D (FLAIR sequence, ET label) | 0.88 [0.45, 1.15] | 0.51 [0.004, 27.53] |  |  |  |  |
|  | szm_lgze_3D (FLAIR sequence, TC label) | 0.93 [0.69, 1.35] | 1.23 [0.08, 14.69] |  |  |  |  |
|  | stat_skew (T1 sequence, TC label) | 0.86 [0.65, 1.07] | 1.06 [0.72, 2.47] |  |  |  |  |

The clinical–radiomic model, combining Age and 10 RFs, achieved the best C-index in the discovery dataset (0.67, 95% CI: 0.63–0.70). The model yielded a C-index of 0.71 (95% CI: 0.61–0.79) on the hold-out test dataset, indicating good prognostic performance (Table 4). Notably, two specific RFs emerged as strong predictors of high-risk status, each exhibiting a Hazard Ratio (HR) greater than 2.0. Szm_lgze_3D, obtained from FLAIR sequence data and corresponding to ET label, revealed an HR of 2.46 (95% CI: [0.004–27.53]). Simultaneously, Morph_pca_flatness, extracted from the TC label, demonstrated an HR of 2.20 (95% CI: [1.00–6.30]). The texture feature szm_lgze_3D captures the frequency and extent of zones with low grey-level intensities within the ET region, providing insight into lesion heterogeneity. Morph_pca_flatness, a shape-based feature, represents the ratio of the least principal axis length and the major principal axis length. The metric’s value exhibits convergence toward one as the TC label geometrically approximates a perfect spherical configuration, thus representing enhanced shape uniformity. In the external validation dataset, the clinical-radiomic model achieved a C-index of 0.64.

**Table 4.** Multivariate Cox regression analysis for Clinical-Radiomic Model.

|  |  | Discovery cohort |  | Holdout test cohort |  | Discovery cohort |  | Holdout test cohort |  |
| --- | --- | --- | --- | --- | --- | --- | --- | --- | --- |
| Model | Variable | HR [95% CI] |  | HR [95% CI] |  | p-Value | C-Index | p-Value | C-Index |
| Clinical-Radiomic Model | Age | 1.36 | [1.17-1.67] | 1.92 | [1.23-4.29] | $1 \times 10^{-8}$ | 0.67 [0.63-0.70] | $2 \times 10^{-4}$ | 0.71 [0.61-0.79] |
|  | morph_pca_maj_axis (TC label) | 1.23 | [0.89-1.67] | 1.72 | [0.68-9.95] |  |  |  |  |
|  | morph_pca_flatness (TC label) | 1.17 | [0.81, 1.47] | 2.20 | [1.00- 6.30] |  |  |  |  |
|  | morph_comp_1 (TC label) | 0.90 | [0.58, 1.53] | 1.60 | [0.52- 8.76] |  |  |  |  |
|  | morph_vol_dens_aee (TC label) | 0.94 | [0.58, 1.53] | 0.36 | [0.08- 1.91] |  |  |  |  |
|  | morph_area_dens_aee (TC label) | 1.04 | [0.74, 1.59] | 1.98 | [0.48-8.57] |  |  |  |  |
|  | ngl_dc_entr_3D (FLAIR sequence, WT label) | 0.87 | [0.70, 1.04] | 0.54 | [0.16- 1.17] |  |  |  |  |
|  | dzm_zdnu_3D (FLAIR sequence, TC label) | 1.14 | [0.88, 1.73] | 1.07 | [0.37-2.18] |  |  |  |  |
|  | szm_lgze_3D (FLAIR sequence, ET label) | 0.85 | [0.42, 1.12] | 2.46 | [0.004-27.53] |  |  |  |  |
|  | szm_lgze_3D (FLAIR sequence, TC label) | 0.99 | [0.74, 1.38] | 0.54 | [0.08, 14.69] |  |  |  |  |
|  | stat_skew (T1 sequence, TC label) | 0.88 | [0.67, 1.11] | 0.92 | [0.55, 2.20] |  |  |  |  |

The log-rank test assessed survival differences between low- and high-risk groups based on a Kaplan–Meier cut-off of 0.012 (Supplementary Materials, Table S-3). The model demonstrated significant stratification ability, revealing distinct survival outcomes between predicted risk groups in the training (discovery), hold-out test, and external validation datasets (p = 1 × 10⁻⁸, p = 2 × 10⁻⁴, and p = 0.01, respectively; Figure 4a,b,c. The Kaplan-Meier survival plots showed the model’s consistent ability to distinguish between high- and low-risk patient groups across datasets. Statistical analysis confirmed significant differences between risk groups, which were also evident in the external validation cohort.

**Figure 4.**
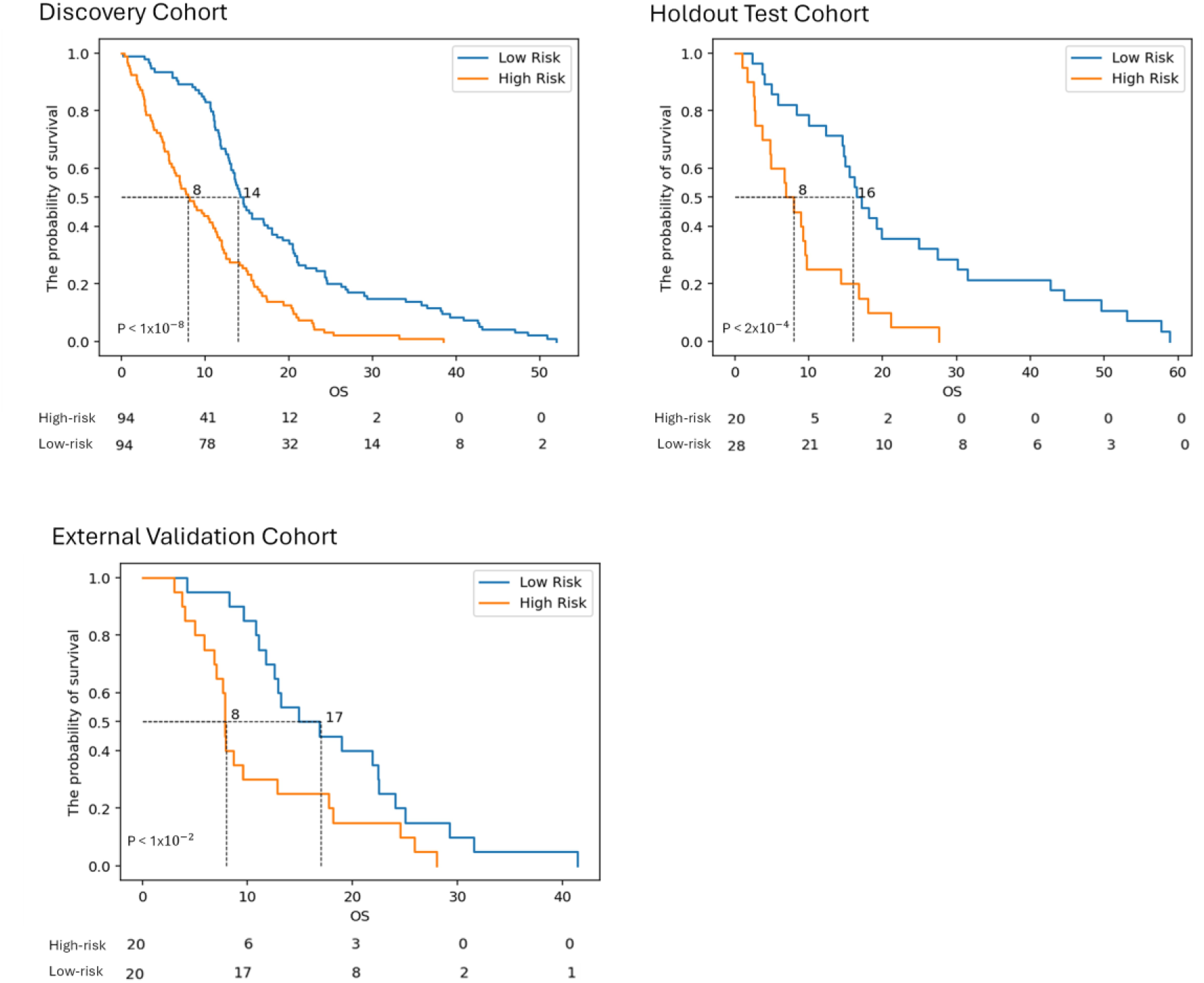
Kaplan–Meier curves display survival differences in the (a) training (discovery), (b) hold-out test and (c) external validation datasets, categorized into low- and high-risk groups by the Cox–LASSO model. The small p-values suggest strong statistical reliability in distinguishing between risk groups.

The permutation importance for each feature is shown in Figure 5 with further information such as feature weights and the Kaplan–Meier threshold presented in Supplementary Materials, Table S-3. According to feature importance analysis based on the final model’s weights, the clinical feature, age, emerged as the most influential predictor overall. Among the RFs specifically, morph_pca_maj_axis was identified as the most significant contributor to the model’s output. This morphological feature evaluates the maximum axial dimension of the ROI-defining ellipsoid for the TC label, determined via principal component analysis and represented by the major eigenvalue (λ_major_). Analytical results indicated that both patient age and the morph_pca_maj_axis feature significantly elevated the likelihood of assignment to the high-risk patient group. The collective effect of these factors indicates that older age and an increased major axis length of the ellipsoid encompassing the ROI may serve as significant predictors of poor prognosis and increased tumor aggressiveness.

**Figure 5.**
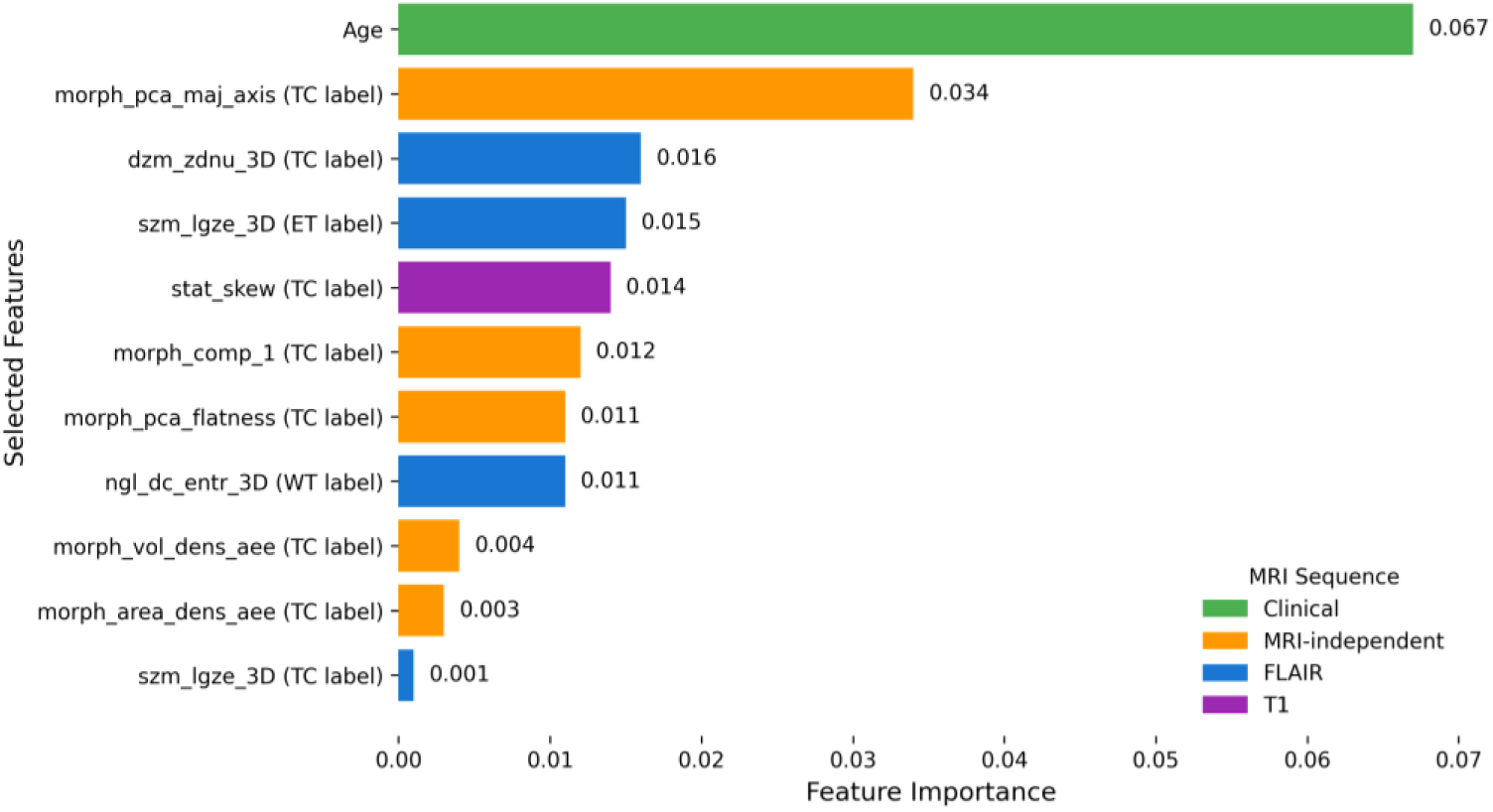
The feature importance for the final clinical-radiomic model.

## 4. Discussion

Utilizing preoperative MRI, this research develops a risk stratification model that includes both a clinical factor and RFs, facilitating the differentiation of GBM patients into distinct low- and high-risk categories. Ten RFs were selected using the LASSO-PSO selection method, which was differentially derived from FLAIR and T1 MRI sequences. The analysis showed that morphological features, shape-based, formed the majority of the selected feature set (5/10 RFs), while texture-based features extracted from FLAIR sequences constituted the second-largest component (4/10 RFs). T1 provided just one first-order feature, while all RFs, except two texture features derived from ET and WT labels, were obtained from the TC label. In the hold-out test dataset, the clinical–radiomic model achieved a C-index of 0.71, demonstrating effective and statistically significant differentiation between risk groups. External validation further supported its robustness, with a C-index of 0.64 and a significant log-rank test result confirming its ability to generalize.

Table 5 provided a comparison of the findings from this study with those from earlier research. Only studies using RFs (engineered or deep features) and clinical data were included. Excluded from the analysis were studies that utilized subjective measures (e.g., VASARI) or RFs lacking reproducibility standards prior to the initial IBSI study [26]. The comparison was further restricted to studies addressing GBM and applying time-to-event survival analysis for OS prediction. A detailed evaluation of the methodological approaches determined important limitations, particularly in single-center studies, potentially decreasing their applicability across heterogeneous clinical settings. Fathi et al. [13] showed standard single-center constraints, while Gomaa et al.’s study [11] enhanced their study’s impact through external validation. We addressed study limitations by expanding the study cohort, incorporating two datasets sourced from multiple centers; utilizing BraTS 2020 for internal analysis and RHUH-GBM for external validation protocols. While this approach improves the study’s reliability, potential biases may still exist, highlighting the need for additional validation in diverse clinical settings.

**Table 5.** The comparison of the proposed study with recent radiomics studies.

| Study | ML Model | Log-rank Test Significance | C-index (Dataset) | Feature Details | IBSI 1,2 Standardized | Feature Limitations (3-10 features) |
| --- | --- | --- | --- | --- | --- | --- |
| Verduin et al. [12] | Multi model Cox regression | Yes | 0.69 (Maastricht UMC + and Radboudumc) | 5 RFs (engineered), 5 Clinical Features (age, sex, EOR, Adjuvant treatment, MGMT) | Partially (not aligned with IBSI 2) | Yes |
| Fathi et al. [13] | Cox-PH | Yes | 0.7 (UPenn) | 24 RFs (deep features, engineered features), 3 Clinical Features (age, sex, EOR) | No | No |
| Duman et al. [38] | Cox-LASSO | Yes | 0.69 (BraTS 2020+ STORM_GLIO) | 2 RFs (engineered), 1 Clinical feature | Yes | Yes |
| Gomaa et al. [11] | Transformer-based DL model | Yes | 0.71(UPenn), 67 (UCSF), 62 (RHUH-GBM) | No info for number of Deep features, 4 Clinical Features (age, sex, MGMT, kps) | No | No |
| Al-Tashi et al. [15] | SwarmDeepSurv (SI-based DeepSurv) | No (0.14>0.05) | 0.61 (TCGA-GBM) | 49 RFs, Clinical Feature (Not provided) | Yes | No |
| Proposed | Cox-LASSO | Yes | 0.71 BraTS 2020, 0.64 (RHUH-GBM) | 10 RFs (engineered), 1 Clinical Feature (Age) | Yes | No |
| Proposed (10 limit) | Cox-LASSO | Yes | 0.71 BraTS 2020, 0.63 (RHUH-GBM) | 7 RFs (engineered), 1 Clinical Features (Age) | Yes | Yes |

Verduin et al. [12] yielded a C-index of 0.69 using five radiomic and five clinical features. In contrast, Duman et al. [38] achieved the same performance with only three features (2 RFs and a single clinical feature). Notably, both studies lacked open-access external validation. The study conducted by Fathi et al. [13] achieved a C-index of 0.70, employing a model consisting of 24 RFs and three clinical variables. Additionally, Gomaa et al. [11] reported C-indexes of 0.71, 0.67, and 0.62 across the UPenn, UCSF, and RHUH-GBM cohorts, respectively, utilizing numerous deep features and four clinical variables. Similarly, Verduin et al. [12] reported a C-index of 0.69 using convolutional filters, which did not comply with IBSI guidelines [39]. The radiomic model developed by Al-Tashi et al. [15], which combined SI algorithms, DL-based survival analysis, and 49 RFs, demonstrated modest predictive capacity (C-index: 0.61) and failed to achieve statistical significance (p = 0.14). In contrast to the common use of Cox regression in radiomics studies [35], Al-Tashi et al. and Gomaa et al. examined SwarmDeepSurv and transformer-based DL architectures for survival analysis. Although these methods demonstrate potential, their lack of transparency presents considerable challenges for clinical integration.

Using only 10 RFs from FLAIR and T1 sequences and the clinical feature (age), our study achieved C-indices of 0.71 (BraTS, 2020) and 0.64 (RHUH-GBM), matching or surpassing prior studies while relying on a minimal feature set. This study found that our method, utilizing an interpretable Cox-LASSO model and reproducible RFs compliant with IBSI standards [26], yielded superior performance metrics when assessed using identical external validation data (RHUH-GBM) compared to the study by Gomaa et al. [11]. Our novel SI-based method, LASSO-PSO, identified 10 RFs, including one clinical feature. The morphological feature morph_pca_maj_axis demonstrated the most significant impact within the validation set. The model demonstrated strong cross-institutional utility through optimized feature selection, maintaining predictive accuracy across varied clinical settings and multiple sourced datasets. The assessment of feature weights and importance (shown in Figure 5 and Supplementary Table S-3) suggests that high-risk patients exhibit an increased axis length (the major eigenvector extracted from principal component analysis), defined by Morph_pca_maj_axis, a morphological feature. Derived from the Gray Level Distance Zone Matrix (GLDZM), Dzm_zdnu_3D feature, Zone Distance Non-Uniformity (ZDNU), computes the degree of uniformity in zone distribution at varying distances, providing insight into the spatial heterogeneity of the tumor. Higher values of the feature indicated greater tumor heterogeneity, which was associated with an increased likelihood of aggressive tumor types and assignment to the high-risk patient category. The TC label was utilized to quantify the two most significant RFs. Within this feature subset, dzm_zdnu_3D was extracted from FLAIR MRI sequences, highlighting its important role in the characterization of tumor aggressiveness. LASSO-PSO surpassed LASSO-RANK and LASSO-GA in selecting an optimal feature subset for radiomic modeling in this study. This result aligns with the comprehensive review of Rostami et al. [29] of SI-based feature selection methods, which show the PSO algorithm’s ability to select a minimum number of features without compromising model performance compared to other SI algorithms.

To the best of our knowledge, this is the first study to apply an SI-based feature selection with traditional ML models that achieves statistically significant risk stratification in time-to-event analysis. Despite utilizing a limited set of clinical and RFs, the proposed model achieved performance levels that were superior to previous studies. Following the guideline set by van Timmeren et al. [10], we excluded RFs demonstrating the low feature importance (<0.01), as shown in Figure 5, from the initial set of 10 RFs. The excluded features were Dzm_zdnu_3D, derived from FLAIR sequence data using the TC label, as well as Morph_vol_dens_aee and Morph_area_dens_aee, both of which were extracted using the TC label. This methodological approach resulted in a finalized feature set of 8 features, which included age as a clinical variable. This revised model preserved discriminative power, with C-indices of 0.71 (BraTS 2020) and 0.63 (RHUH-GBM). Although the constraint limiting the analysis to feature subsets of 3–10 features significantly impacted the features evaluated, the potential for the SI-based selection method to enhance performance if applied to an expanded set of RFs represents a promising avenue for future research.

In medical research, the data scarcity and imbalance due to rare conditions, small sample sizes, and incomplete clinical records limit the availability of robust datasets [40]. To enhance clinical applicability and reduce data dependency, this study adopted an approach by incorporating only one clinical variable (age) and utilizing only two MRI sequences (FLAIR and T1) instead of all four MRI sequences. Potential retrospective sampling bias was systematically mitigated through the utilization of data derived from multiple institutions (the BraTS 2020 dataset), with further evaluation provided with external validation procedures (RHUH-GBM dataset). In the future, model optimization could cover integrating expanded clinical parameters (genetic markers, MGMT, KPS etc.), additional imaging modalities (PET, CT, ultrasound), and other omics datasets (genomics, pathomics). In order to prioritize reproducibility and interpretability, which are still challenging to clinical implementation, we deliberately exclude DL-based RFs. The deep features can enhance survival prediction performance, addressing the current limitations in future studies [41]. Following the IBSI standardization of convolutional filters [39], future studies could focus on implementing these standardized filters. Additionally, we aim to investigate the utility of SI-based approaches for model development, specifically exploring their potential to improve predictive performance.

## 5. Conclusion

This study presents an interpretable clinical–radiomic model for the prediction of OS, developed and validated using a novel SI-based hybrid feature selection approach to stratify GBM patients into low- and high-risk categories. In accordance with established radiomic research guidelines, the presented study implemented a methodology integrating SI-based hybrid feature selection with a Cox-LASSO. Furthermore, the research managed important clinical challenges while enhancing interpretability and minimizing model complexity, highlighting its potential for clinical integration and reproducibility across various clinical settings. This framework performance is comparable to or exceeds the predictive results recently reported by Poursaeed et al. [35] based on the previously determined inclusion criteria. Expanding radiomic studies to contain additional data sources and validating them with larger, multi-institutional cohorts can potentially improve its predictive power, thereby may help the transition of radiomic studies into clinical decision-making in the treatment of GBM.

## Data Availability

All data produced in the present study are available upon reasonable request to the authors.

https://www.cancerimagingarchive.net/collection/rhuh-gbm/

https://www.kaggle.com/datasets/awsaf49/brats2020-training-data

## 7 Supplementary Materials

**Figure-S6.**
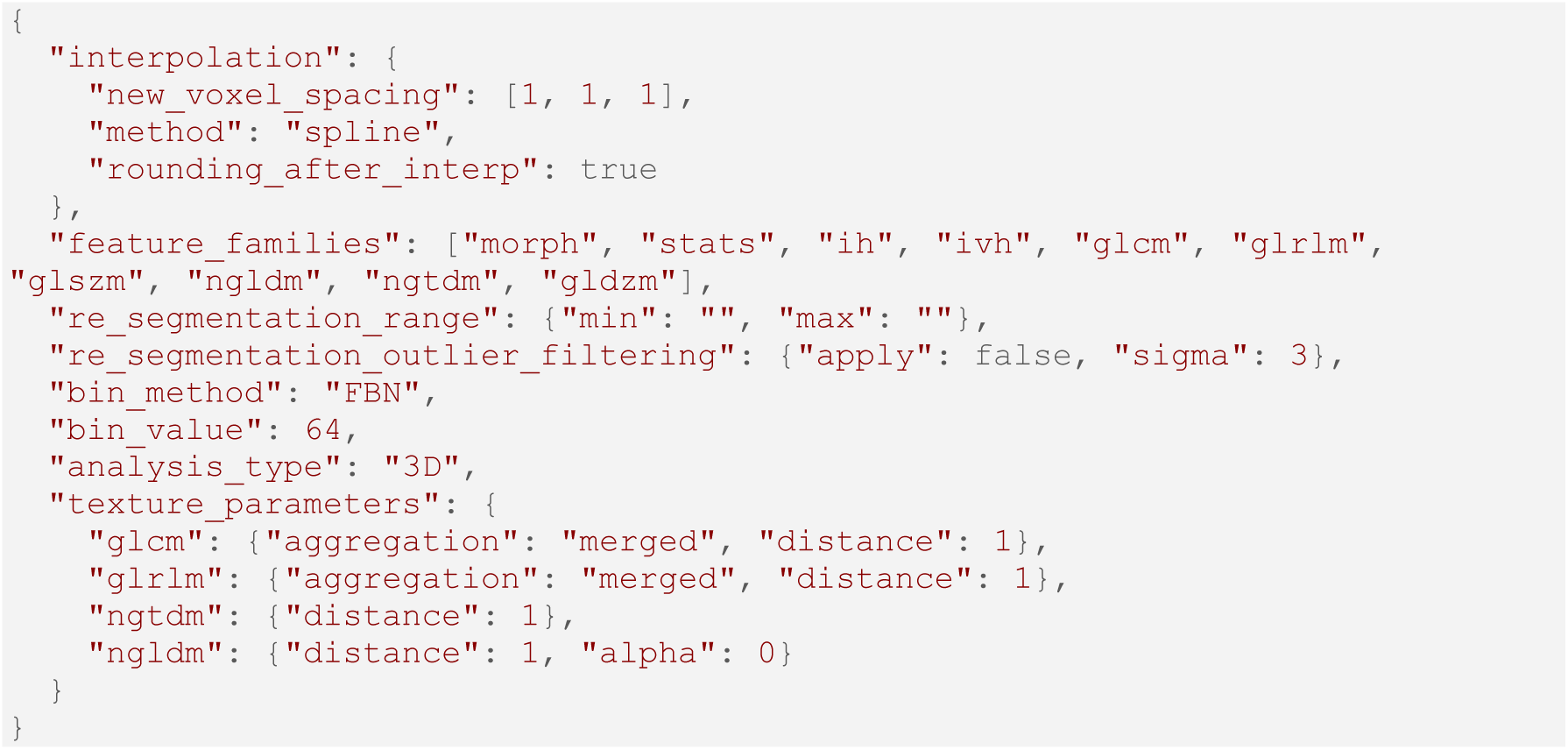
The IBSI standardized preprocessing parameters for radiomic analysis.

**Figure-S7.**
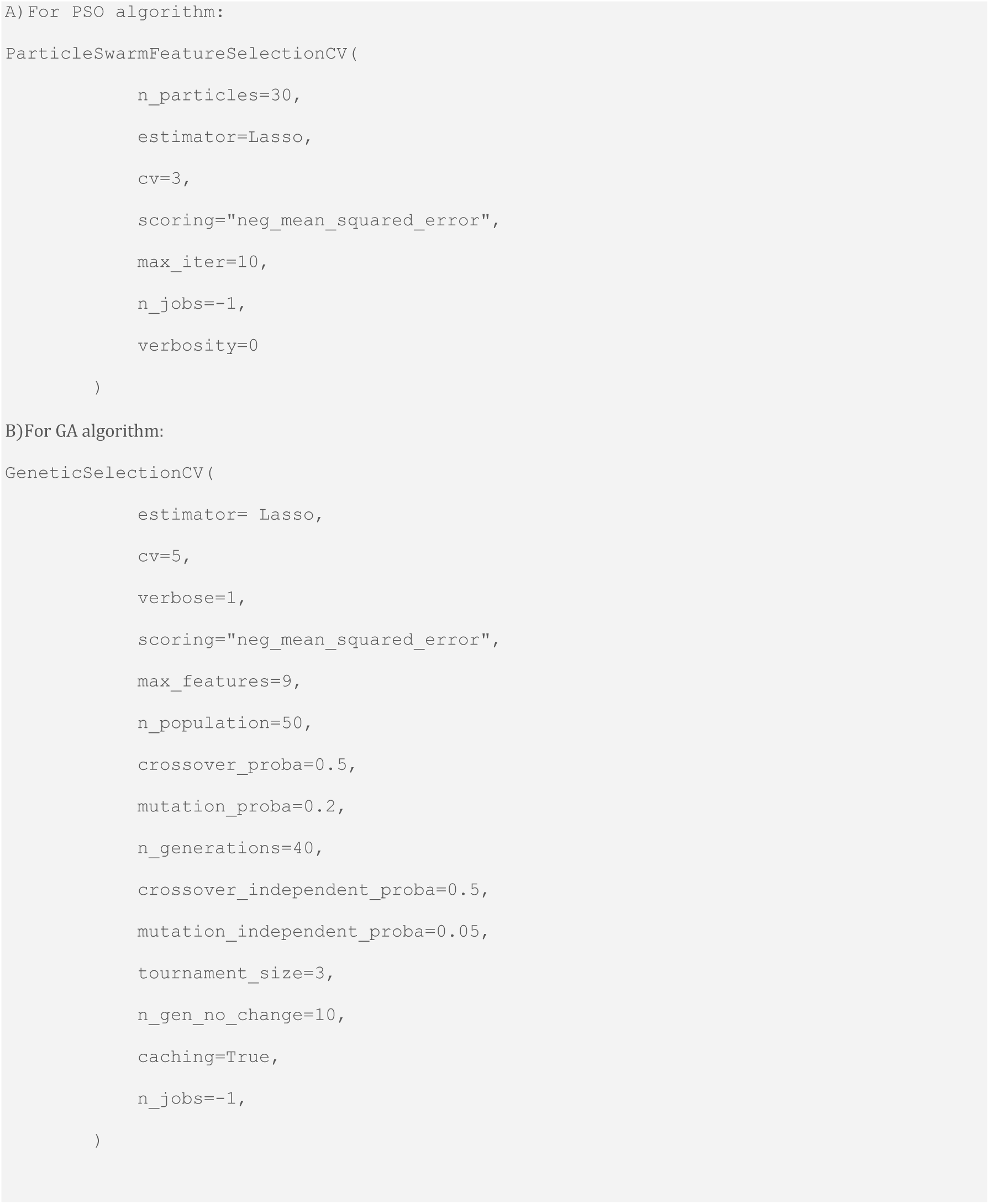
Hyperparameters for (A) PSO and (B) GA were set based on the example source codes, with the exception of ‘max_features’, which was adjusted to 9 (the total feature number for each feature pool) for GA, leading to improved model performance.

**Figure-S8.**
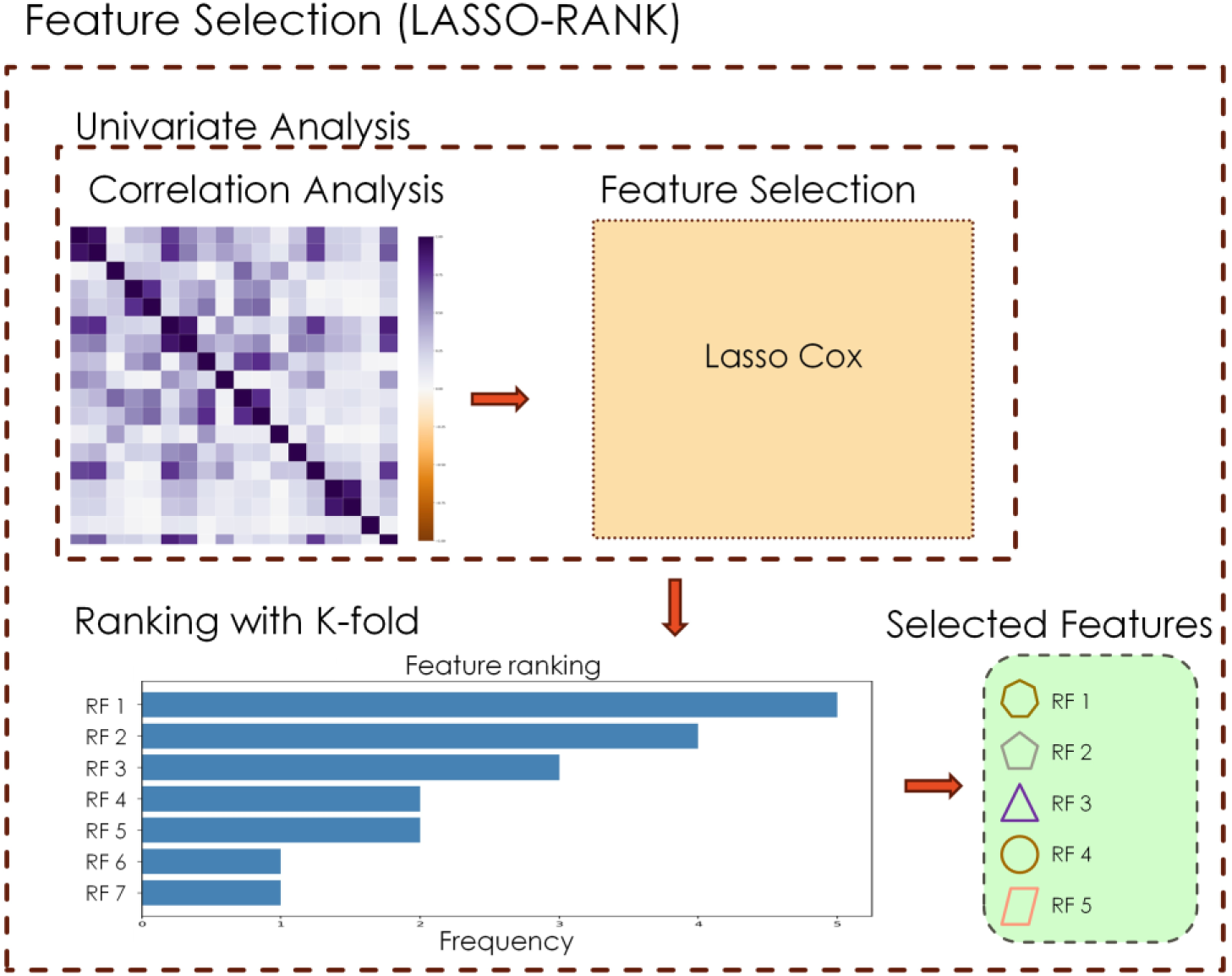
The LASSO-RANK feature selection framework

**Figure-S9.**
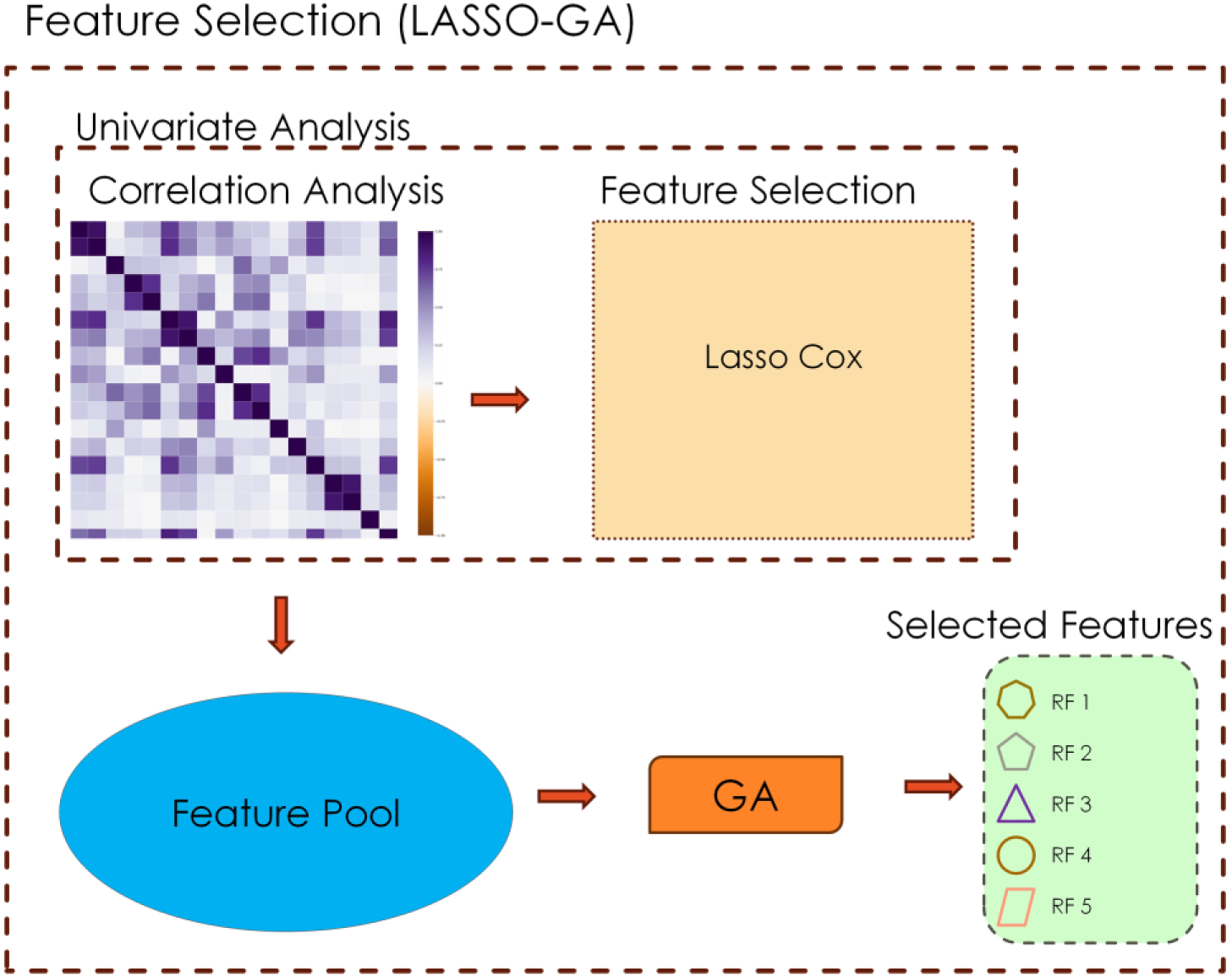
The LASSO-GA feature selection framework.

**Figure-S10.**
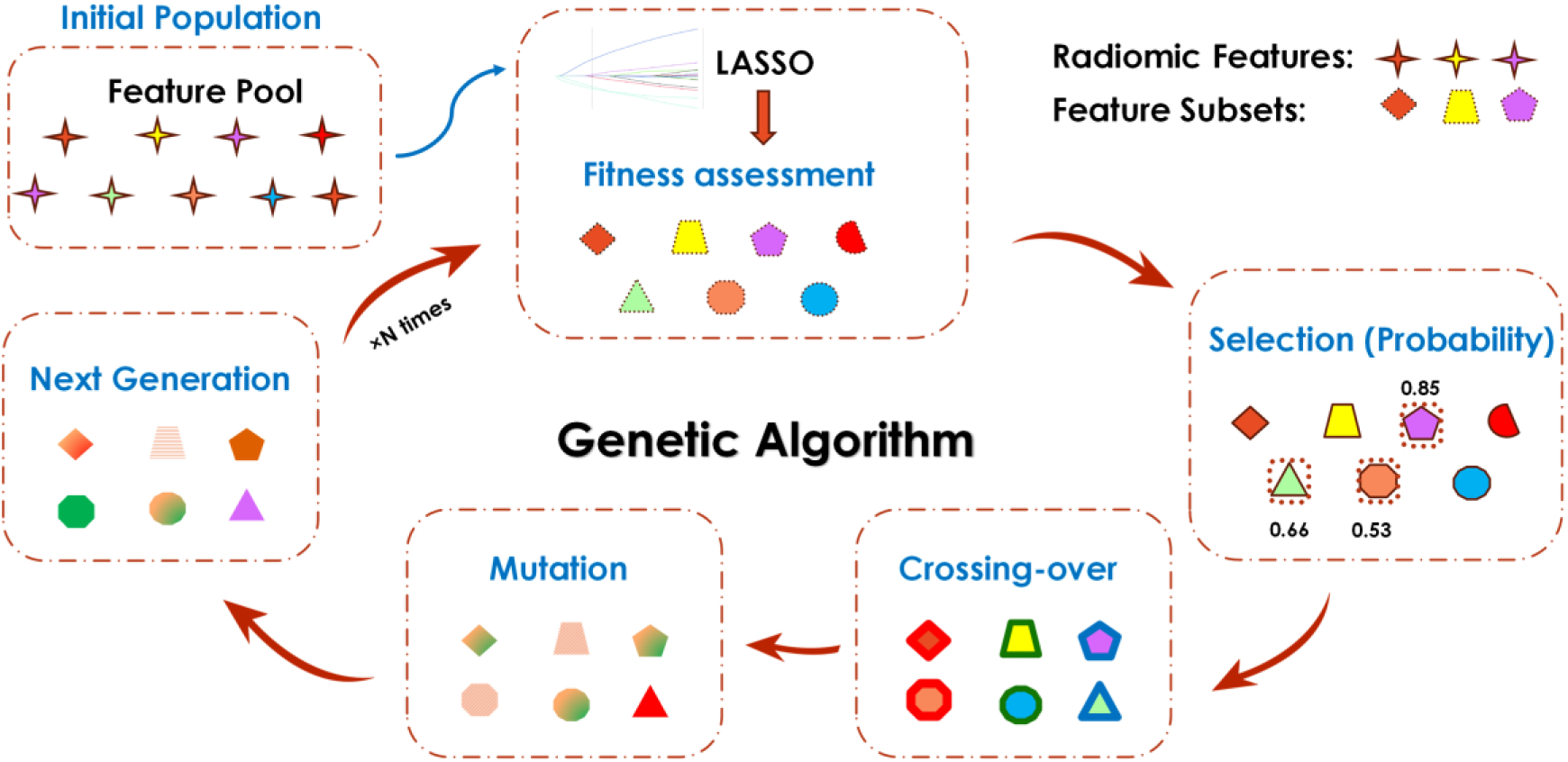
GA-based Feature Selection Workflow with the feature pool from LASSO. RFs: radiomic features, FS

**Figure-S11.**
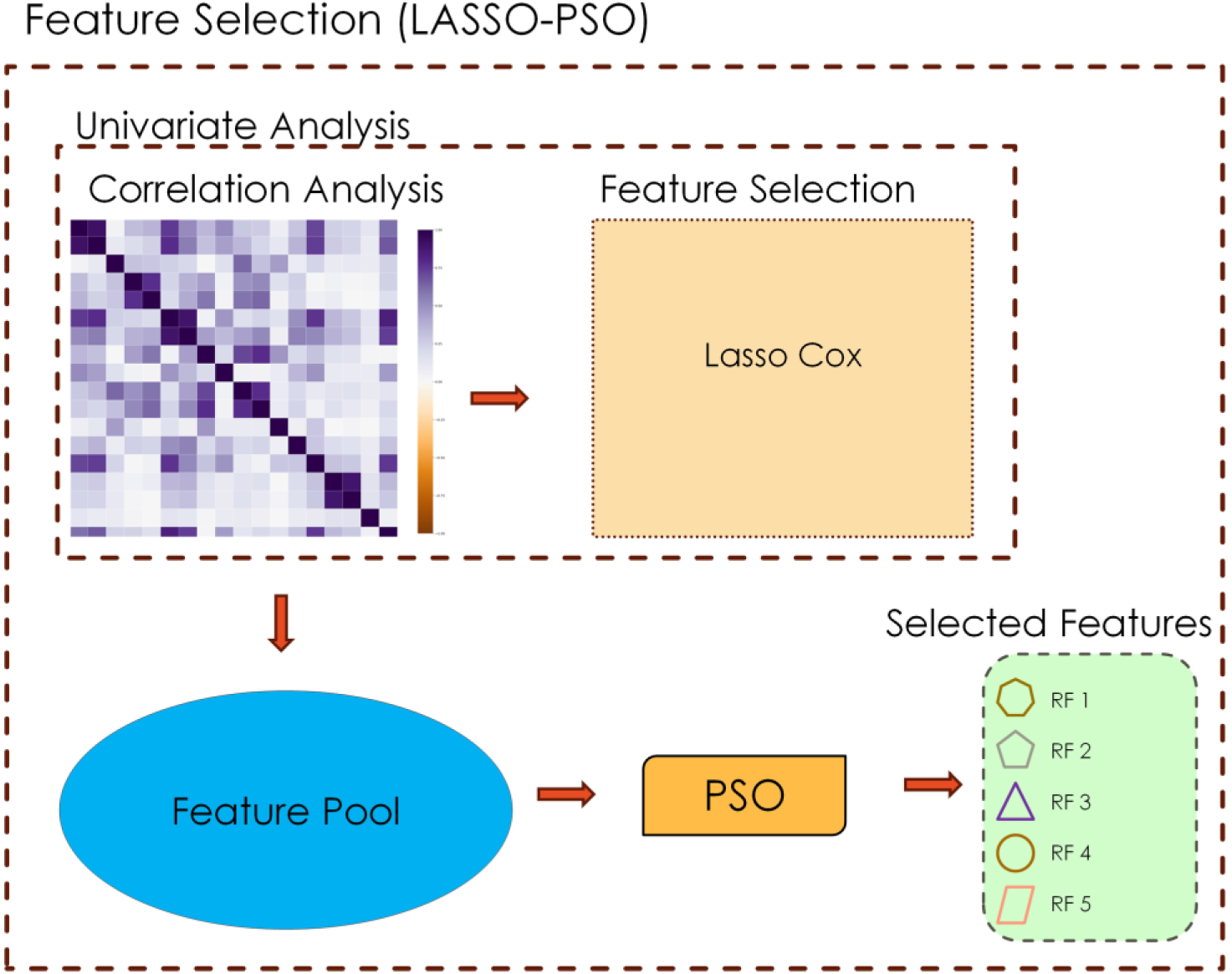
The LASSO-PSO feature selection framework

**Figure-S12.**
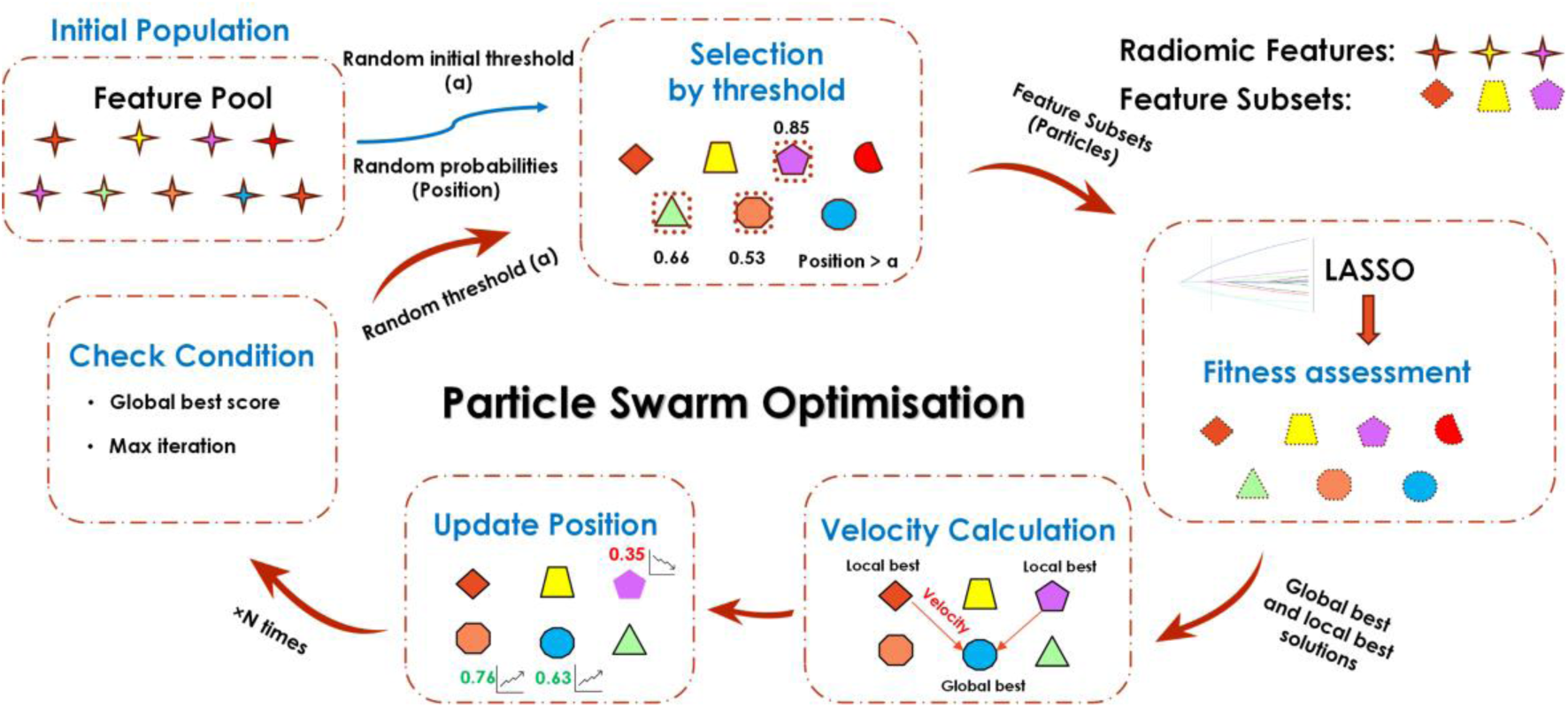
PSO-based feature selection workflow with the feature pool from LASSO

**Table-S6.** The hyperparameters for each model and feature selection method from 200 bootstrapped iterations of the training (discovery) dataset.

| Model | Feature Selection | Hyperparameters<br>(selected [min, max]) |
| --- | --- | --- |
| Cox-Lasso | LASSO-RANK | alpha = 50 [1,100] |
| Cox-Lasso | LASSO-GA | alpha = 100 [1,100] |
| Cox-Lasso | LASSO-PSO | alpha = 5 [1,100] |
| RSF | LASSO-RANK | n_estimators = 10 [3,10]<br>max_depth = 5 [3,5]<br>min_samples_split = 3 [3,10] |
| RSF | LASSO-GA | n_estimators = 5 [3,10]<br>max_depth = 5 [3,5]<br>min_samples_split = 10 [3,10] |
| RSF | LASSO-PSO | n_estimators = 10 [3,10]<br>max_depth = 4 [3,5]<br>min_samples_split = 10 [3,10] |

**Table-S7.** The RSF model performance on the discovery, the hold-out test, and the external validation.

|  |  | C-Index |  |  |
| --- | --- | --- | --- | --- |
| Model | Feature Selection Method | Discovery Cohort | Hold-out Test Cohort | External Validation Cohort |
| RSF | LASSO-PSO | 0.74 | 0.63 | 0.58 |

**Table-S8.**
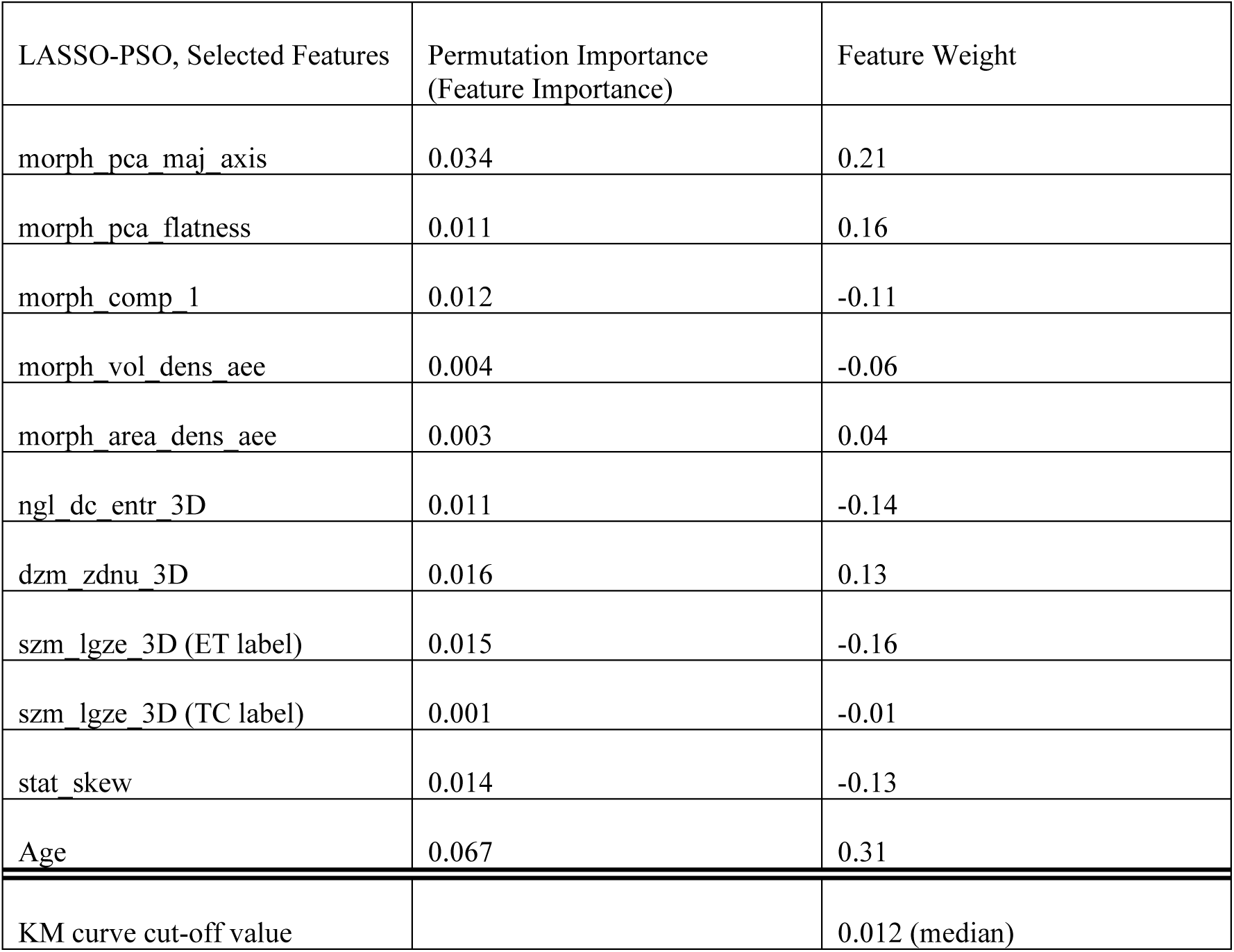
The feature Impartance and weights of each feature in the final clinical-radiomic mode.

## References

[1] K. D. Miller et al., “Brain and other central nervous system tumor statistics, 2021,” CA Cancer J Clin, vol. 71, no. 5, pp. 381–406, 2021.

[2] M. A. Qazi et al., “Intratumoral heterogeneity: pathways to treatment resistance and relapse in human glioblastoma,” Annals of Oncology, vol. 28, no. 7, pp. 1448–1456, 2017, doi: 10.1093/annonc/mdx169.

[3] D. N. Louis et al., “The 2021 WHO Classification of Tumors of the Central Nervous System: a summary,” Neuro Oncol, vol. 23, no. 8, pp. 1231–1251, Aug. 2021, doi: 10.1093/neuonc/noab106.

[4] W. Wu et al., “Glioblastoma multiforme (GBM): An overview of current therapies and mechanisms of resistance,” Pharmacol Res, vol. 171, p. 105780, 2021.

[5] R. J. Jackson et al., “Limitations of stereotactic biopsy in the initial management of gliomas,” Neuro Oncol, vol. 3, no. 3, pp. 193–200, 2001.

[6] A. G. Rockall, A. Hatrick, P. Armstrong, and M. Wastie, Diagnostic Imaging. Hoboken, UNITED KINGDOM: John Wiley & Sons, Incorporated, 2013. [Online]. Available: http://ebookcentral.proquest.com/lib/cardiff/detail.action?docID=1154368

[7] P. Lambin et al., “Radiomics: the bridge between medical imaging and personalized medicine,” Nat Rev Clin Oncol, vol. 14, no. 12, pp. 749–762, 2017.

[8] R. J. Gillies, P. E. Kinahan, and H. Hricak, “Radiomics: Images Are More than Pictures, They Are Data,” Radiology, vol. 278, no. 2, pp. 563–577, Nov. 2015, doi: 10.1148/radiol.2015151169.

[9] A. Rabasco Meneghetti et al., “Definition and validation of a radiomics signature for loco-regional tumour control in patients with locally advanced head and neck squamous cell carcinoma,” Clin Transl Radiat Oncol, vol. 26, pp. 62–70, 2021, doi: 10.1016/j.ctro.2020.11.011.

[10] J. E. van Timmeren, D. Cester, S. Tanadini-Lang, H. Alkadhi, and B. Baessler, “Radiomics in medical imaging—‘how-to’ guide and critical reflection,” Insights Imaging, vol. 11, no. 1, p. 91, 2020, doi: 10.1186/s13244-020-00887-2.

[11] A. Gomaa et al., “Comprehensive multimodal deep learning survival prediction enabled by a transformer architecture: A multicenter study in glioblastoma,” Neurooncol Adv, vol. 6, no. 1, p. vdae122, Jan. 2024, doi: 10.1093/noajnl/vdae122.

[12] M. Verduin et al., “Prognostic and predictive value of integrated qualitative and quantitative magnetic resonance imaging analysis in glioblastoma,” Cancers (Basel), vol. 13, no. 4, pp. 1–20, Feb. 2021, doi: 10.3390/cancers13040722.

[13] A. Fathi Kazerooni et al., “Clinical measures, radiomics, and genomics offer synergistic value in AI-based prediction of overall survival in patients with glioblastoma,” Sci Rep, vol. 12, no. 1, p. 8784, 2022, doi: 10.1038/s41598-022-12699-z.

[14] D. T. Do, M.-R. Yang, L. H. T. Lam, N. Q. K. Le, and Y.-W. Wu, “Improving MGMT methylation status prediction of glioblastoma through optimizing radiomics features using genetic algorithm-based machine learning approach,” Sci Rep, vol. 12, no. 1, p. 13412, 2022, doi: 10.1038/s41598-022-17707-w.

[15] Q. Al-Tashi et al., “SwarmDeepSurv: swarm intelligence advances deep survival network for prognostic radiomics signatures in four solid cancers,” Patterns, vol. 4, no. 8, 2023.

[16] A. G. Tzalavra, I. Andreadis, K. V Dalakleidi, F. Constantinidis, E. I Zacharaki, and K. S Nikita, “Dynamic contrast enhanced-magnetic resonance imaging radiomics combined with a hybrid adaptive neuro-fuzzy inference system-particle swarm optimization approach for breast tumour classification,” Expert Syst, vol. 39, no. 4, p. e12895, 2022.

[17] X. Pan, C. Liu, T. Feng, and X. S. Qi, “A multi-objective based radiomics feature selection method for response prediction following radiotherapy,” Phys Med Biol, vol. 68, no. 5, p. 055018, 2023.

[18] B. H. Menze et al., “The Multimodal Brain Tumor Image Segmentation Benchmark (BRATS),” IEEE Trans Med Imaging, vol. 34, no. 10, pp. 1993–2024, 2015, doi: 10.1109/TMI.2014.2377694.

[19] S. Bakas et al., “Advancing the cancer genome atlas glioma MRI collections with expert segmentation labels and radiomic features,” Sci Data, vol. 4, no. 1, pp. 1–13, 2017.

[20] S. Bakas et al., “Identifying the best machine learning algorithms for brain tumor segmentation, progression assessment, and overall survival prediction in the BRATS challenge,” arXiv preprint arXiv:1811.02629, 2018.

[21] S. Cepeda et al., “The Río Hortega University Hospital Glioblastoma dataset: A comprehensive collection of preoperative, early postoperative and recurrence MRI scans (RHUH-GBM),” Data Brief, vol. 50, p. 109617, 2023.

[22] P. Y. Wen, S. M. Chang, M. J. Van den Bent, M. A. Vogelbaum, D. R. Macdonald, and E. Q. Lee, “Response Assessment in Neuro-Oncology Clinical Trials,” Journal of Clinical Oncology, vol. 35, no. 21, pp. 2439–2449, Jun. 2017, doi: 10.1200/JCO.2017.72.7511.

[23] N. J. Tustison et al., “N4ITK: improved N3 bias correction,” IEEE Trans Med Imaging, vol. 29, no. 6, pp. 1310–1320, 2010.

[24] T. Rohlfing, N. M. Zahr, E. V Sullivan, and A. Pfefferbaum, “The SRI24 multichannel atlas of normal adult human brain structure,” Hum Brain Mapp, vol. 31, no. 5, pp. 798–819, 2010.

[25] S. Pati et al., “The Cancer Imaging Phenomics Toolkit (CaPTk): Technical Overview,” in Brainlesion: Glioma, Multiple Sclerosis, Stroke and Traumatic Brain Injuries, A. Crimi and S. Bakas, Eds., Cham: Springer International Publishing, 2020, pp. 380–394.

[26] A. Zwanenburg et al., “The image biomarker standardization initiative: standardized quantitative radiomics for high-throughput image-based phenotyping,” Radiology, vol. 295, no. 2, pp. 328–338, 2020.

[27] P. Whybra, C. Parkinson, K. Foley, J. Staffurth, and E. Spezi, “Assessing radiomic feature robustness to interpolation in 18F-FDG PET imaging,” Sci Rep, vol. 9, no. 1, p. 9649, 2019, doi: 10.1038/s41598-019-46030-0.

[28] C. Piazzese, K. Foley, P. Whybra, C. Hurt, T. Crosby, and E. Spezi, “Discovery of stable and prognostic CT-based radiomic features independent of contrast administration and dimensionality in oesophageal cancer,” PLoS One, vol. 14, no. 11, p. e0225550, 2019, [Online]. Available: 10.1371/journal.pone.0225550

[29] M. Rostami, K. Berahmand, E. Nasiri, and S. Forouzandeh, “Review of swarm intelligence-based feature selection methods,” Eng Appl Artif Intell, vol. 100, p. 104210, 2021.

[30] R. Tibshirani, “The lasso method for variable selection in the Cox model,” Stat Med, vol. 16, no. 4, pp. 385–395, 1997.

[31] S. Leger et al., “A comparative study of machine learning methods for time-to-event survival data for radiomics risk modelling,” Sci Rep, vol. 7, no. 1, p. 13206, 2017, doi: 10.1038/s41598-017-13448-3.

[32] J. H. Holland, “Genetic algorithms,” Sci Am, vol. 267, no. 1, pp. 66–73, 1992.

[33] J. Kennedy and R. Eberhart, “Particle swarm optimization,” in Proceedings of ICNN’95-international conference on neural networks, ieee, 1995, pp. 1942–1948.

[34] H. Ishwaran, U. B. Kogalur, E. H. Blackstone, and M. S. Lauer, “Random survival forests,” Ann Appl Stat, vol. 2, no. 3, pp. 841–860, Sep. 2008, doi: 10.1214/08-AOAS169.

[35] R. Poursaeed, M. Mohammadzadeh, and A. A. Safaei, “Survival prediction of glioblastoma patients using machine learning and deep learning: a systematic review,” BMC Cancer, vol. 24, no. 1, p. 1581, 2024.

[36] O. W. Mwangi, A. Islam, and O. Luke, “Bootstrap Confidence Intervals for Proportions of Unequal Sized Groups Adjusted for Overdispersion,” Open J Stat, vol. 5, no. 6, pp. 502–510, 2015.

[37] J.-C. Luo, Q.-Y. Zhao, and G.-W. Tu, “Clinical prediction models in the precision medicine era: old and new algorithms,” Ann Transl Med, vol. 8, no. 6, 2020.

[38] A. Duman, X. Sun, S. Thomas, J. R. Powell, and E. Spezi, “Reproducible and interpretable machine learning-based radiomic analysis for overall survival prediction in glioblastoma multiforme,” Cancers (Basel), vol. 16, no. 19, p. 3351, 2024.

[39] P. Whybra et al., “The Image Biomarker Standardization Initiative: Standardized Convolutional Filters for Reproducible Radiomics and Enhanced Clinical Insights,” Radiology, vol. 310, no. 2, p. e231319, Feb. 2024, doi: 10.1148/radiol.231319.

[40] F. Mandreoli, D. Ferrari, V. Guidetti, F. Motta, and P. Missier, “Real-world data mining meets clinical practice: Research challenges and perspective,” Front Big Data, vol. 5, 2022, [Online]. Available: https://www.frontiersin.org/journals/big-data/articles/10.3389/fdata.2022.1021621

[41] X. Zhang et al., “Deep Learning With Radiomics for Disease Diagnosis and Treatment: Challenges and Potential,” Front Oncol, vol. 12, 2022, [Online]. Available: https://www.frontiersin.org/articles/10.3389/fonc.2022.773840

